# LST-AI: a Deep Learning Ensemble for Accurate MS Lesion Segmentation

**DOI:** 10.1101/2023.11.23.23298966

**Authors:** Tun Wiltgen, Julian McGinnis, Sarah Schlaeger, Florian Kofler, CuiCi Voon, Achim Berthele, Daria Bischl, Lioba Grundl, Nikolaus Will, Marie Metz, David Schinz, Dominik Sepp, Philipp Prucker, Benita Schmitz-Koep, Claus Zimmer, Bjoern Menze, Daniel Rueckert, Bernhard Hemmer, Jan Kirschke, Mark Mühlau, Benedikt Wiestler

**Author notes:** Corresponding Author: Mark Mühlau Department of Neurology, School of Medicine, Technical University of Munich Ismaninger Str. 22, 81675 Munich, Germany. indicates equal contribution (Mark Mühlau, Benedikt Wiestler).

## Abstract

Automated segmentation of brain white matter lesions is crucial for both clinical assessment and scientific research in multiple sclerosis (MS). Over a decade ago, we introduced an engineered lesion segmentation tool, LST. While recent lesion segmentation approaches have leveraged artificial intelligence (AI), they often remain proprietary and difficult to adopt. As an open-source tool, we present LST-AI, an advanced deep learning-based extension of LST that consists of an ensemble of three 3D-UNets.

LST-AI explicitly addresses the imbalance between white matter (WM) lesions and non-lesioned WM. It employs a composite loss function incorporating binary cross-entropy and Tversky loss to improve segmentation of the highly heterogeneous MS lesions. We train the network ensemble on 491 MS pairs of T1w and FLAIR images, collected in-house from a 3T MRI scanner, and expert neuroradiologists manually segmented the utilized lesion maps for training. LST-AI additionally includes a lesion location annotation tool, labeling lesion location according to the 2017 McDonald criteria (periventricular, infratentorial, juxtacortical, subcortical). We conduct evaluations on 103 test cases consisting of publicly available data using the Anima segmentation validation tools and compare LST-AI with several publicly available lesion segmentation models.

Our empirical analysis shows that LST-AI achieves superior performance compared to existing methods. Its Dice and F1 scores exceeded 0.62, outperforming LST, SAMSEG (Sequence Adaptive Multimodal SEGmentation), and the popular nnUNet framework, which all scored below 0.56. Notably, LST-AI demonstrated exceptional performance on the MSSEG-1 challenge dataset, an international WM lesion segmentation challenge, with a Dice score of 0.65 and an F1 score of 0.63—surpassing all other competing models at the time of the challenge. With increasing lesion volume, the lesion detection rate rapidly increased with a detection rate of >75% for lesions with a volume between 10mm^3^ and 100mm^3^.

Given its higher segmentation performance, we recommend that research groups currently using LST transition to LST-AI. To facilitate broad adoption, we are releasing LST-AI as an open-source model, available as a command-line tool, dockerized container, or Python script, enabling diverse applications across multiple platforms.

## 1. Introduction

Multiple sclerosis (MS) is a complex chronic inflammatory disease of the central nervous system. Clinically, MS typically manifests through neurological deficits which are mainly driven by inflammatory demyelinating lesions occurring in brain white matter and in the spinal cord and by neurodegeneration (axonal and neuronal loss). To date, inflammatory white matter lesions are a hallmark of MS and their identification on magnetic resonance imaging (MRI) plays a crucial role in the diagnosis and follow-up of MS (Filippi et al., 2018; Thompson, Banwell, et al., 2018; Thompson, Baranzini, et al., 2018). In addition, the location of lesions within the brain plays a role in diagnosing MS, as lesions in periventricular, juxtacortical, and infratentorial regions are part of the MS diagnostic criteria by indicating dissemination in space. In contrast, lesions in the subcortical region are only used to determine longitudinal dissemination and to monitor disease progression (Thompson, Banwell, et al., 2018).

In clinical routine and research, the gold standard of lesion identification and segmentation is manual segmentation by trained neuroradiological experts. However, this constitutes a time-consuming task with both relevant inter- and intra-rater variability, thereby hampering studies with large datasets aiming to improve our understanding of MS.

In past years, many algorithms and tools have been developed and published to accurately automate lesion segmentation. As one of the early contributions to this field, we published the Lesion Segmentation Toolbox (LST), which has since been applied in numerous scholarly publications (Schmidt et al., 2012). While early segmentation algorithms have been designed primarily using statistical and early machine learning models such as Support Vector Machines, Gaussian Mixture Models or engineered by using manually selected features (Schmidt et al., 2012), more recent approaches incorporate learning-based features via encoder/decoder model stages (Cerri et al., 2021) or learn these end to end in fully convolutional models in (semi-) supervised settings (Commowick et al., 2018). With the advent of artificial intelligence (AI), automated lesion segmentation tools based on convolutional neural networks (CNN) have become increasingly popular and indeed provide similar or higher segmentation accuracy than earlier, machine learning-based methods (Diaz-Hurtado et al., 2022; H. Li et al., 2018; Ma et al., 2022; Zeng et al., 2020). This is also reflected in the rankings of published MS lesion segmentation challenges, e.g., MICCAI 2016 (Commowick et al., 2018) and ISBI 2015 (Carass et al., 2017). While CNN-based models often outperform earlier models in challenges, they only excel with a sufficient number of training data, as they are designed to learn priors and features automatically and do not incorporate manual feature selection. Consequently, they are especially prone to overfitting to the training data. Moreover, and in contrast to earlier machine learning models, CNNs are comparatively harder to regularize, as they have higher model and learning capacity, larger number of model parameters and thus more complex loss landscapes. Therefore, a large performance gap between training set and test set is often noticeable and highlights the need to evaluate the performance of CNN-based models on heterogeneous external test data. Overcoming this gap and generalizing segmentation models in order to be applicable to data from multiple protocols and centers is one of the main on-going challenges for AI-based approaches. In this context, some AI-based approaches that have previously been published are optimized towards transferability: Valverde et al. have provided *nicMSlesions*, a CNN-based lesion segmentation method that is able to adjust to a new image domain by retraining their model on a single image (Valverde et al., 2019). An important CNN-based architecture is the UNet, which has been applied in many previous lesion segmentation studies (Ashtari et al., 2022; Hashemi et al., 2022; Krishnan et al., 2023; La Rosa et al., 2020; Ronneberger et al., 2015). Furthermore, recent studies successfully train their models on one dataset and test it on another, external dataset, for which the MICCAI 2016 (Commowick et al., 2021) and ISBI 2015 (Carass et al., 2017) datasets are often selected (Cerri et al., 2021; Gentile et al., 2023; Kamraoui et al., 2022; Krishnan et al., 2023; X. Li et al., 2022; McKinley et al., 2021). Hence, the research field is moving towards more generalized segmentation tools, which is an important step towards clinical applicability of these methods.

In this study, we introduce a deep learning-based extension of LST. The main contributions can be outlined in three aspects: 1) We provide an open-source lesion segmentation tool (with network weights) that is easy to use and maintained; 2) The tool has been validated on external datasets; 3) Lesion segmentation performance is comparable to or better than state of the art. We carefully explain our selection of model architecture and describe the training and test set used, and show how our composite loss function allows us to optimize our model for generalizability on MRIs of unseen test centers. We also compare the performance of our model against existing MS lesion segmentation algorithms. To facilitate studies and applications in MS research, we provide this enhanced toolkit as open source to the imaging community (https://github.com/CompImg/LST-AI).

## 2. Methods

### 2.1. Datasets

In the following section, we characterize and define training and test set, including details on image acquisition. With regard to in-house data, we respected the Code of Ethics of the World Medical Association (Declaration of Helsinki) for experiments involving humans; the study was approved by the local ethics committee.

For the training set, we used an in-house dataset consisting of 491 paired 3D FLAIR and 3D T1w images acquired on a 3.0T Achieva scanner (Philips Medical Systems, Best, The Netherlands) to train both our proposed LST-AI segmentation model and the nnUNet baseline. Testing and evaluation of segmentation performance of all methods was conducted on multiple datasets. The test set includes four publicly available datasets: (i) msisbi: ISBI 2015 training data (Carass et al., 2017) (https://smart-stats-tools.org/lesion-challenge-2015); (ii) msljub: dataset published by Laboratory of Imaging Technologies (Lesjak et al., 2018) (https://lit.fe.uni-lj.si/en/research/resources/3D-MR-MS/); (iii) mssegtest: MICCAI 2016 challenge test dataset (Commowick et al., 2021) (https://shanoir.irisa.fr/shanoir-ng/welcome) and (iv) mssegtrain: MICCAI 2016 challenge training dataset (Commowick et al., 2021) (https://shanoir.irisa.fr/shanoir-ng/welcome). One case (msseg-test-center07-08) was removed from the mssegtest dataset because it included incorrect ground truth data. In total, the test set consists of 103 images from 87 subjects (note that the publicly available ISBI dataset is a longitudinal dataset). Further characteristics of the datasets, including data on lesion load, are provided in Table 1. Details on image acquisition are provided in Table 2.

**Table 1.**
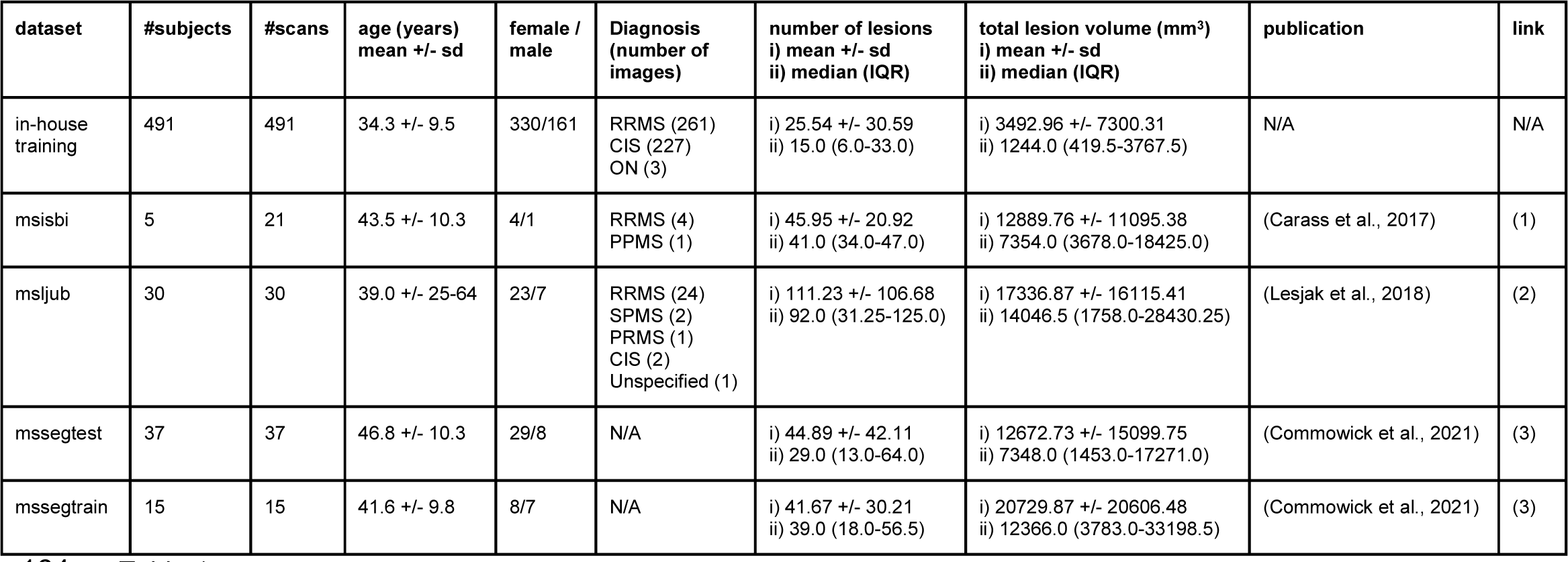
Characteristics of the datasets. One in-house (training) dataset was used, as well as the public datasets msisbi from the ISBI 2015 challenge (Carass et al., 2017), msljub published by the Laboratory of Imaging Technologies (Lesjak et al., 2018), and mssegtest and mssegtrain which are the testing and training datasets from the MICCAI 2016 challenge, respectively (Commowick et al., 2021). Abbreviations: CIS: clinically isolated syndrome, IQR: interquartile range, N/A: not applicable/available, ON: optic neuritis, PPMS: primary progressive multiple sclerosis, RRMS: relapsing-remitting multiple sclerosis, sd: standard deviation, SPMS: secondary progressive multiple sclerosis (1) https://smart-stats-tools.org/lesion-challenge-2015 (2) https://lit.fe.uni-lj.si/en/research/resources/3D-MR-MS/ (3) https://shanoir.irisa.fr/shanoir-ng/welcome

**Table 2.**
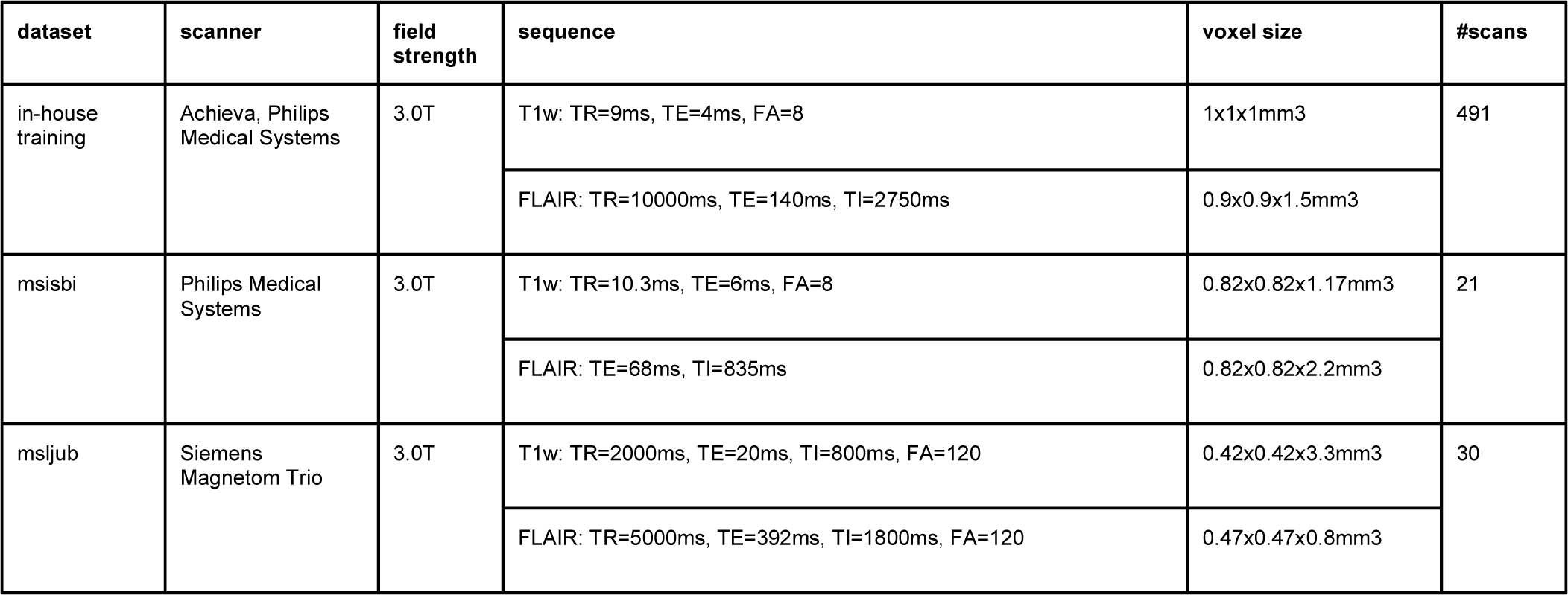
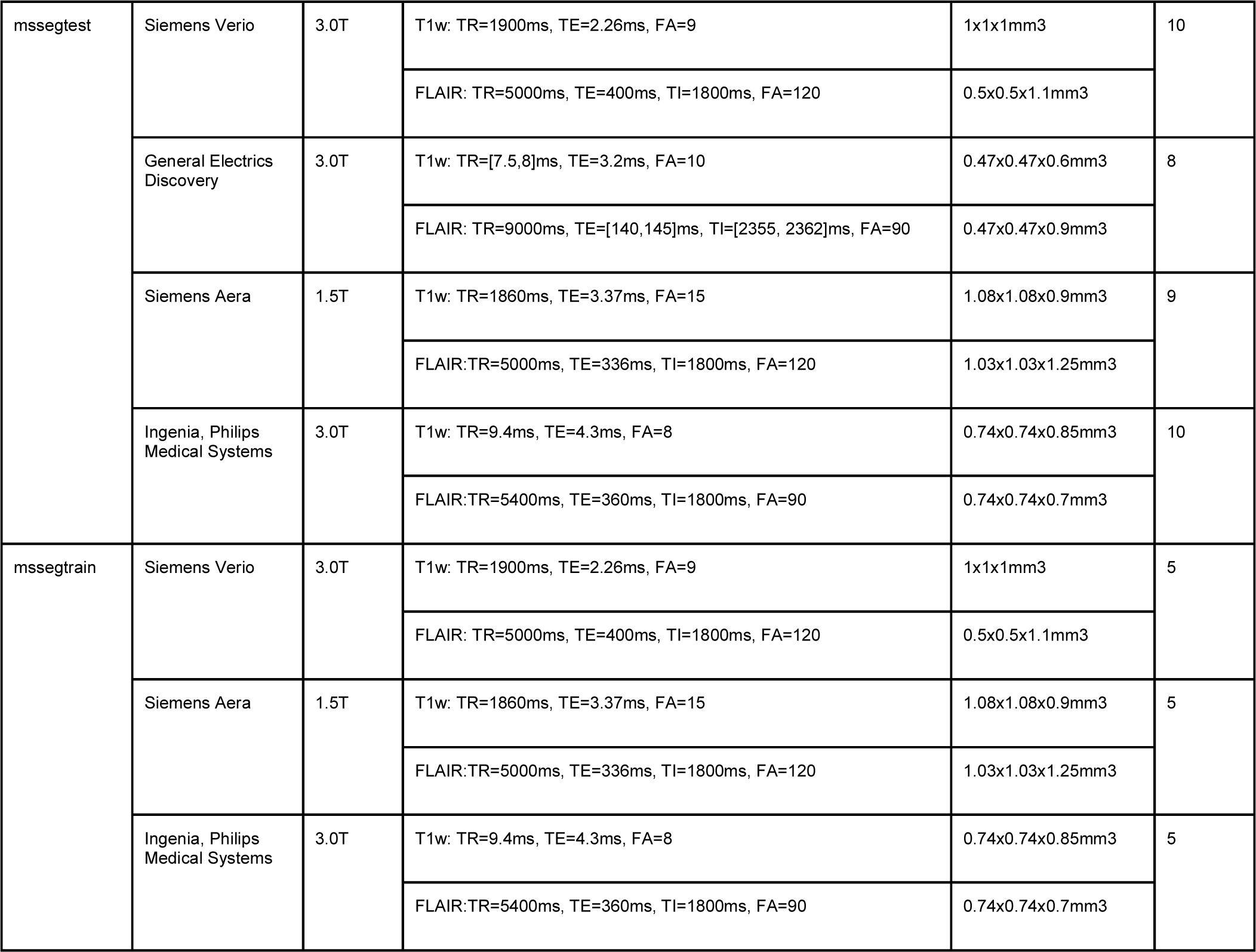
Acquisition settings of the datasets. Abbreviations: FA: flip angle, FLAIR: fluid-attenuated inversion recovery, TE: echo time, TI: inversion time, TR: repetition time, T1w: T1-weighted

### 2.2 Preprocessing

To guarantee fair comparisons across all baselines, we standardize preprocessing across all datasets and methods. Firstly, we register (rigid registration) all images to the MNI ICBM152 nonlinear atlas version 2009 template (https://www.mcgill.ca/bic/neuroinformatics/brain-atlases-human) using the Greedy command line tool (P. Yushkevich, 2016/2023; P. A. Yushkevich et al., 2016). This atlas registration both ensures a consistent voxel resolution (1×1×1mm^3^) and image orientation, preprocessing steps well established for deep learning segmentation models (Kofler et al., 2020; Pati et al., 2022). Subsequently, we use the deep learning-based HD-BET brain extraction tool to generate skull-stripped images (Isensee et al., 2019). Next, the shape of the skull-stripped images is cropped to the size that is required for the 3D UNets and intensities are normalized to [0;1]. To benchmark methods in its intended environment, we opt for non-skull-stripped images for SAMSEG, as well as the legacy algorithms of LST, the Lesion Prediction Algorithm (LST-LPA) and the Lesion Growth Algorithm (LST-LGA), which perform optimally with whole-brain data. Consequently, we omit the HD-BET skull-stripping, cropping, and intensity normalization preprocessing steps for these specific baselines while retaining them for others.

This standardized preprocessing (including skull-stripping) is also integrated into our LST-AI toolbox, providing users with a streamlined approach.

### 2.3. Lesion segmentation

In this section, we first describe the proposed lesion segmentation tool followed by benchmark methods that have been applied in many studies and to which the proposed tool is compared. Finally, we outline the manual lesion segmentation workflows employed across the different datasets.

#### 2.3.1. LST-AI ensemble network

The LST-AI tool encompasses preprocessing, lesion segmentation and, optionally, lesion location annotation. An overview of the workflow is shown in Figure 1.

**Figure 1.**
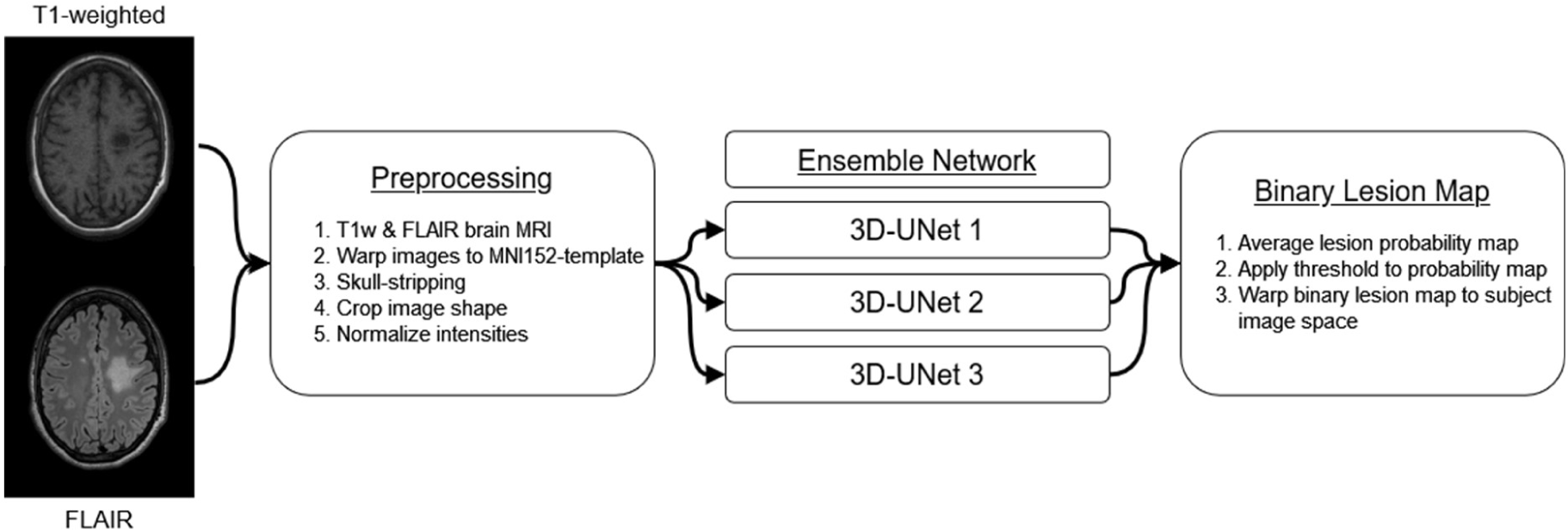
The different processing steps of the holistic LST-AI tool are presented. First, a pair of T1w and FLAIR images is warped to MNI space, then skull-stripped, cropped, and intensity-normalized during preprocessing. The resulting images are used as input for the three 3D-UNets of the ensemble network. Each UNet provides a lesion probability map. To generate the binary lesion map, the three lesion probability maps are averaged and a threshold is subsequently applied. Finally, the binary lesion map is warped back to the subject image space (original space of the FLAIR image).

The preprocessing functionality included in LST-AI is outlined in section 2.2. Specifically, the T1w and FLAIR images are warped to the MNI ICBM152 template, then skull-stripped, center cropped to shape (192, 192, 192), and, finally, intensities were normalized to [0;1]. With respect to the model architecture, LST-AI is based on an ensemble of three 3D-UNets. Each UNet is built upon the 3D-UNet (Çiçek et al., 2016) architecture and inspired by nnUNet (Isensee et al., 2021). It is composed of 5 encoder and 5 decoder blocks. Each of these blocks is built from two convolution blocks (3D convolution, instance normalization, leaky ReLU activation) and skip connections between respective encoder and decoder blocks (see Figure 2). In encoder blocks, downsampling is implemented via strided convolutions with stride 2, while transposed convolutions are used for upscaling in decoder blocks. Following the architectural choices in nnUNet (Isensee et al., 2021), we employ deep supervision layers in the training with the intuition of allowing gradients to flow deeper into the networks’ layers (Wang et al., 2015). The number of deep supervision layers differed for the three UNets: one UNet included one deep supervision layer and the two other UNets included two deep supervision layers to allow for some variability in the ensemble predictions. For the loss function, we used a combination of Tversky loss (Salehi et al., 2017) (with higher penalization of false-negative lesion omissions) and binary cross-entropy in the deep supervision layers and a combined dice loss and binary cross-entropy in the full-resolution output. During training, we randomly chained intensity (random Gaussian noise, random Gaussian smoothing, random gamma adjustment) and geometry augmentations (random flips and crops). Each model was trained for a total of 1000 epochs, using the stochastic gradient descent optimizer (with Nesterov momentum) and a polynomial learning rate decay, starting at 1e-2. This training scheme has been adapted from nnUNet and was shown to generalize well in the medical segmentation decathlon (Antonelli et al., 2022). In total, three training runs were started from scratch to create an ensemble of three models, a technique previously reported (H. Li et al., 2018).

**Figure 2.**
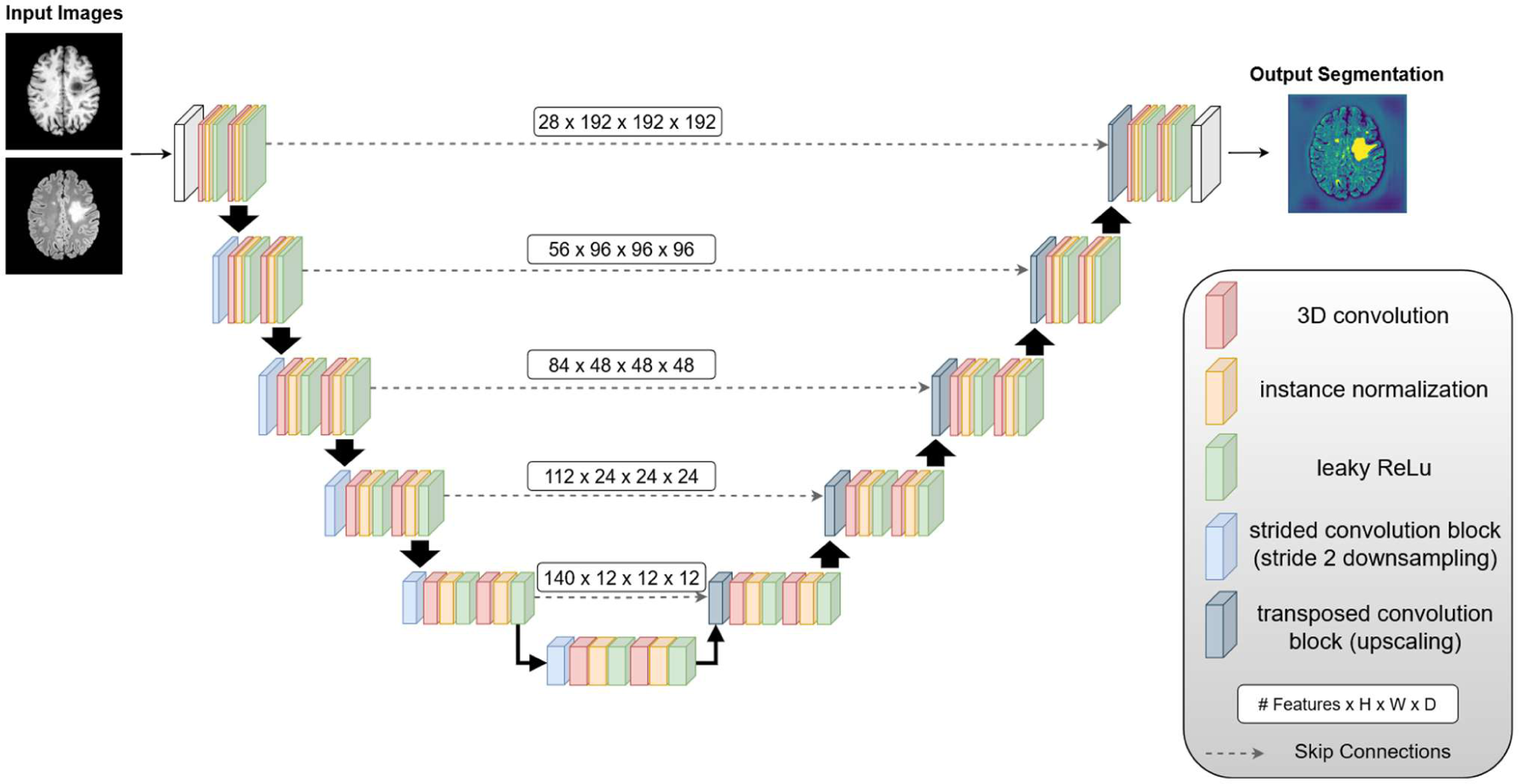
Architecture of the 3D-UNets which constitute the ensemble network of LST-AI. They comprise two channels (one for T1w images and one for FLAIR images) and consist of 5 encoder and 5 decoder blocks. Strided convolutions (stride 2) are used for downsampling and transposed convolutions are used for upscaling. Encoder and decoder blocks are connected via skip connections.

For the final segmentation output, the preprocessed T1w and FLAIR images are used as input for each one of the 3D UNets which generate three lesion probability maps. The final binary lesion map is obtained by averaging the three lesion probability maps and subsequent thresholding (default threshold of 0.5). This workflow, including the ground truth lesion segmentation mask, is illustrated in Figure 3, using an example of the msljub dataset (subject 05).

**Figure 3.**
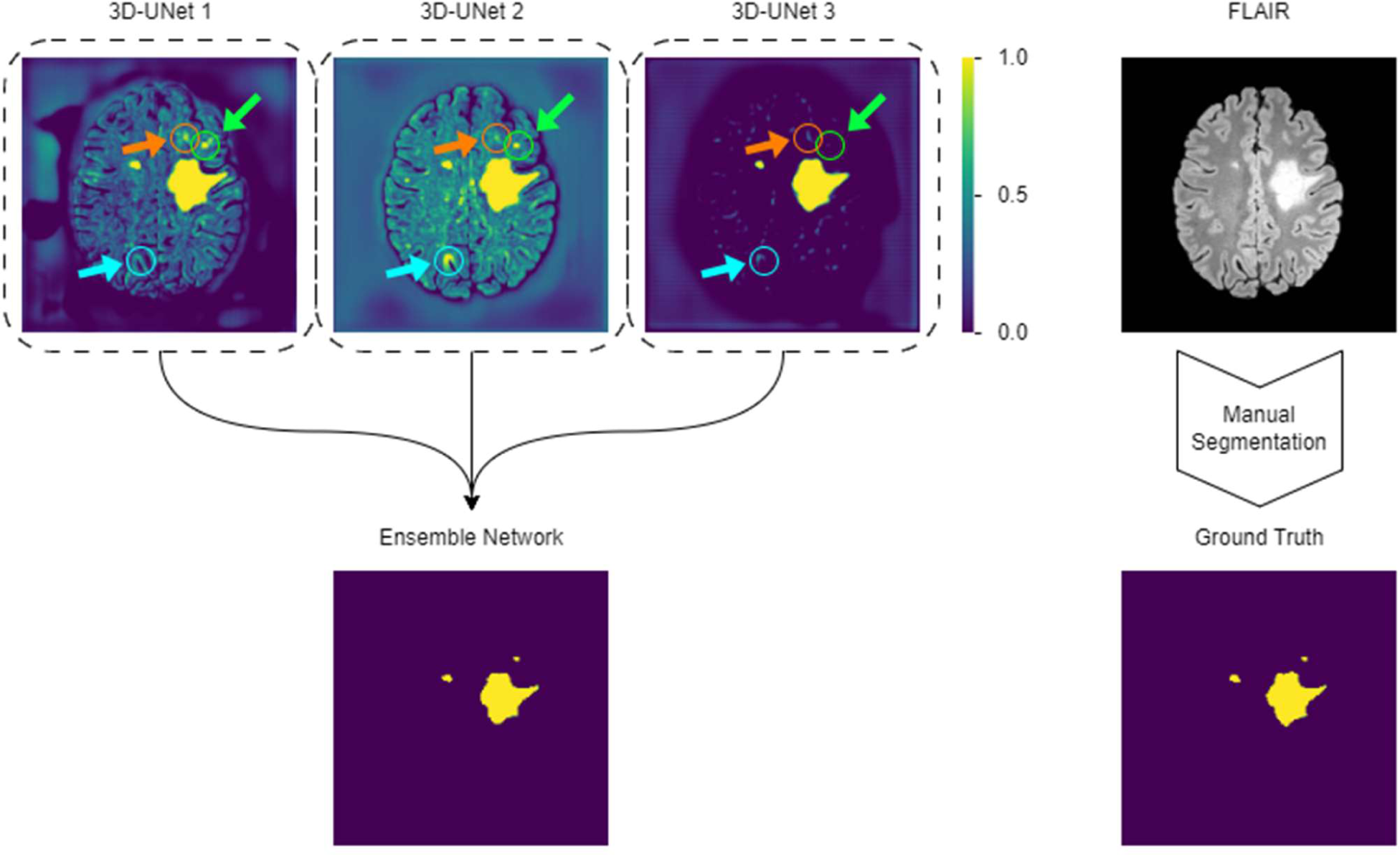
Rationale behind the ensemble network of LST-AI. First, the three 3D-UNets generate a lesion probability map. The mean of the three outputs is calculated and thresholded to generate the final binary lesion map. On the right-hand side, we show a slice of a FLAIR image and the corresponding manual segmentation (i.e., the ground truth). The orange arrow and circle highlight a false positive present in the lesion probability map of 3D-UNet 1, but not in the other lesion probability maps. The light blue arrow and circle highlight a false positive present in the lesion probability map of 3D-UNet 2, but not in the other lesion probability maps. The green arrow and circle highlight a false negative lesion in the lesion probability map of 3D-UNet 3, which is detected by 3D-UNet 1 and 2. Note how the output of the ensemble network is more accurate than the output of the individual networks, as it does not show the false positives and false negatives.

As an additional feature, the tool can optionally label lesions according to their location, i.e., periventricular (PV), juxtacortical (JC), subcortical (SC), or infratentorial (IT). To this end, the same MNI ICBM152 nonlinear T1 atlas used above is first registered deformably (using Greedy) to the skull-stripped T1w image in MNI space. The resulting transformation is applied to a manually labeled anatomical mask indicating different brain regions (inter alia: ventricles for PV labeling, infratentorial region for IT labeling, cortex for JC labeling, and subcortical region for SC labeling), which is thereby registered to the skull-stripped T1w image in MNI space. The anatomical mask is shown in Figure 4. Next, each individual lesion from the binary lesion segmentation map is dilated using a cube as footprint (3×3×3mm^3^), and assigned to the region with which it overlaps by at least one voxel (e.g., if a dilated lesion overlaps with the ventricles of the anatomical mask it is labeled as PV). During this step, lesions are checked to overlap with the four brain regions sequentially so that each lesion can be attributed to only one category. The order of checks is PV, IT, JC, and, finally, SC. By this choice, large lesions overlapping with the inner ventricles and the cortical ribbon are classified as PV (as PV lesions are commonly the largest). In the resulting lesion map, the lesions are labeled according to their location (PV: label=1, JC: label=2, SC: label=3, IT: label=4). Finally, the labeled lesion map is transformed to the original space of the FLAIR image with the inverse of the affine transformation, which was computed earlier, resulting in location-annotated lesion maps in the original subject space as well as in the MNI space.

**Figure 4.**
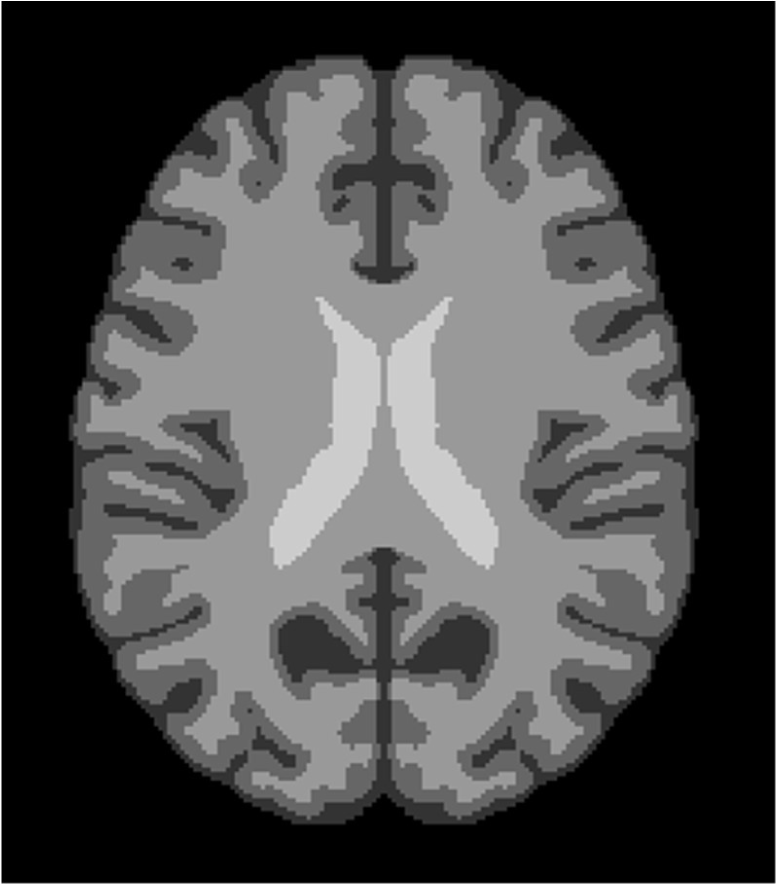
MS-specific anatomical mask indicating four different brain regions: ventricles outlined in light gray (used to label lesions as periventricular), cortex outlined in dark gray (used to label lesions as juxtacortical), subcortical region outlined in gray (used to label lesions as subcortical), or infratentorial region (not visible in the image). Note that lesions are dilated using a 3×3×3mm^3^ cube before overlaying with the anatomical mask, which is how lesions can overlap with ventricle or cortex regions, resulting in lesions labeled as periventricular or juxtacortical, respectively.

We intend to target a diverse user base and provide LST-AI as a set of standalone command line tools and as a dockerized application, including all model checkpoints and required preprocessing tools (Greedy and HD-BET). As LST-AI can be used in similar ways as Freesurfer/FSL command line tools or nicMSlesions (docker), we give the opportunity to conveniently integrate our tool into existing workflows.

For accelerated performance, we recommend using our tool in a GPU-enabled environment but we also provide a fallback method for CPU-only usage. Depending on the exact hardware setup, typical execution time varies between tens of seconds (GPU) and 1-2 minutes on a CPU-only system. We provide LST-AI’s functionality for three different workflows: segmentation-only, lesion location annotation-only, or both. Moreover, labels can be exported in the original subject space or in the MNI ICBM152 template space.

Moreover, we make our source code available, allowing the community to adapt and tailor our tools for different application scenarios, by modifying preprocessing tools or using the checkpoints for pre-training of custom models. We intend to continuously maintain and update our tool in the github repository. In conclusion, while we have high confidence in the generalization capabilities of LST-AI, we want to emphasize that it is explicitly designed for research and non-clinical purposes. It has not undergone the necessary certification or licensing for clinical applications.

#### 2.3.2. Benchmark methods

Evaluation of the performance of the proposed tool is realized through comparison to other publicly available lesion segmentation methods. This includes the widely used LST version 3.0.0 (https://www.applied-statistics.de/lst.html) with its lesion growth algorithm (LGA) (Schmidt et al., 2012) and lesion prediction algorithm (LPA) (Vanderbecq et al., 2020), to which our proposed tool presents a complementary, AI-based lesion segmentation method. Additionally, a trained nnUNet and the recently published SAMSEG lesion segmentation tool implemented in Freesurfer version 7.3.2 (Cerri et al., 2021) are used for comparison.

● **LST-LGA** (Schmidt et al., 2012): This method requires T1w and FLAIR images that are not skull-stripped. Before applying the LST-LGA tool, T1w and FLAIR images are preprocessed as described in section 2.2. Additionally, images are denoised using the CAT12 (Gaser et al., 2022) denoising filter implemented in SPM12 (https://www.fil.ion.ucl.ac.uk/spm/software/spm12/). Then, the LST-LGA lesion segmentation algorithm is applied. First, using the methods implemented in SPM12, bias field correction is applied to the FLAIR image, and the T1w image is segmented into white matter, grey matter, and cerebrospinal fluid. Based on the FLAIR intensities, lesion belief maps are generated for each tissue class. The lesion belief map of grey matter is then thresholded (default threshold of 0.3 as suggested in Schmidt et al., 2012), which results in seeds that are used for the lesion growth model. Thereby, lesion seeds are expanded according to FLAIR hyperintensities, eventually producing a lesion probability map. Finally, a binary lesion map is generated after thresholding the lesion probability map (threshold of 0.5).
● **LST-LPA** (Vanderbecq et al., 2020): This method requires only FLAIR images that are not skull-stripped. Preprocessing is identical to the LST-LGA workflow and includes registration to MNI and denoising. Similarly, bias correction is applied, and a lesion belief map is generated based on FLAIR intensities. The LST-LPA algorithm is a binary regression model that combines the lesion belief map and fixed parameters, which had been learned through logistic regression during the development of the tool in order to calculate the lesion probability map. The binary lesion map is again generated by applying a threshold to the lesion probability map (threshold of 0.5).
● **nnUNet** (Isensee et al., 2021): The UNet’s early achievements in deep learning for biomedical segmentation have led to extensive research in refining its architecture for specialized tasks. Building on this, Isensee et al. (2021) have introduced an innovative framework that automates the selection of hyperparameters and data augmentation techniques based on the specific dataset employed. To provide this baseline, we format our training set according to nn-UNet’s convention and train the model for 1000 epochs with five-fold cross-validation. We select the stronger 3D-UNet baseline in contrast to a 2D-UNet baseline, and use the full-resolution model as a baseline.
● **SAMSEG** (Cerri et al., 2021): This method, Sequence Adaptive Multimodal SEGmentation, requires only one MRI contrast image but it also accepts multiple contrasts. Here, we use T1w and FLAIR image pairs that are not skull-stripped as input. As recommended by the authors (Cerri et al., 2021), preprocessing is minimal, with images only being registered to MNI space using Greedy (P. A. Yushkevich et al., 2016). During the segmentation process, a deformable probabilistic atlas is used as segmentation prior and is iteratively fitted to the input data. Thereby, voxels are assigned to the brain structures with highest probability, including lesions. The binary lesion map is obtained by only selecting the voxels with lesion labels and setting all other voxel values to zero.

For region-specific analyses, all binary lesion maps are annotated with the method implemented in the LST-AI tool. In effect, each lesion is labeled according to its location (i.e., PV, JC, IT, or SC).

#### 2.3.3. Manual segmentation

We make use of multiple datasets. Therefore, the workflows of manual segmentation, i.e., generation of ground truth lesion maps, differ. We describe the manual segmentation protocols of the different datasets and refer to the corresponding publications:

● **in-house training:** The training data were first pre-segmented using LST-LGA. Segmented lesions were manually reviewed and, based on FLAIR images, corrected by one out of four experienced neuroradiologists using ITK-SNAP (P. A. Yushkevich et al., 2006). All lesion masks were eventually reviewed by one senior neuroradiologist. The manual lesion segmentation protocol is also described in another publication using the same dataset (Hapfelmeier et al., 2023).
● **msisbi:** All images were manually delineated by two raters. Since no consensus was available, we arbitrarily selected the lesion maps of one of the two raters as ground truth (rater 2). Protocol details have been described in the original publication (Carass et al., 2017).
● **msljub:** All images were delineated by three raters using a semi-automated approach. A consensus segmentation was obtained through revision of the combined lesion maps by all three raters; a detailed protocol is available in the original publication (Lesjak et al., 2018).
● **mssegtest & mssegtrain:** All images were manually delineated by seven raters, from which a consensus was constructed. Details on the protocol and consensus construction are available in the original publication (Commowick et al., 2021).

## 2.4. Evaluation

To assess the effectiveness of the LST-AI lesion segmentation tool, we compare its results with manual segmentations and other available tools in multiple external datasets to evaluate the performance and generalizability. These external sets encompass various acquisition protocols, scanners, and originate from different centers. For consistency, we use images and lesion maps in MNI space. Our evaluation covers lesion segmentation and detection methods, applying a minimum lesion volume threshold of 3mm³ corresponding to 3 MNI-space voxels.

### 2.4.1. Lesion segmentation

Regarding lesion segmentation evaluation, we rely on the animaSegPerfAnalyzer tool from the anima evaluation toolbox (https://anima.irisa.fr/), which was also used in the MICCAI 2016 MS lesion segmentation challenge (Commowick et al., 2018). It requires pairs of ground truth (i.e, manually segmented) and automatically segmented lesion maps. This toolbox computes various metrics to analyze the segmentation performance at both the voxel and lesion level. Regarding voxel-wise analysis, we were interested in the Dice Similarity Coefficient (DSC):

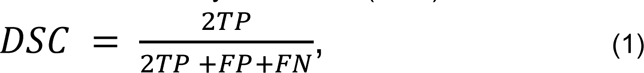

the positive predictive value (PPV):

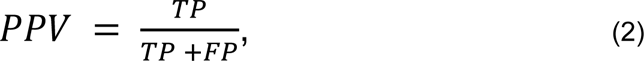

and the sensitivity:

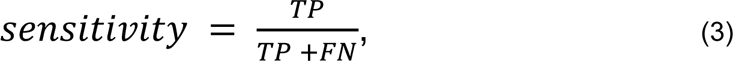

where TP denotes the true positives, FP the false positives, FN the false negatives. In addition, we extracted the average surface distance (ASD) with the animaSegPerfAnalyzer tool:

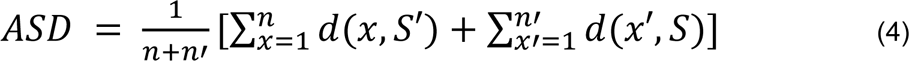

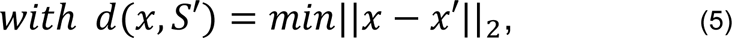

where n and n’ are the number of points x and x’ on the surface S of the manual segmentation and the surface S’ of the automated segmentation, respectively, and d() is the minimal Euclidean distance between a point x on surface S and the surface S′.

These metrics are calculated for each image, then averaged within each dataset, and finally averaged across all datasets. Thereby, we provide an overall score across different scanners and centers as well as individual scores for each dataset.

As an additional step, we construct one array by concatenating all images and calculate the DSC across all lesions of all datasets. We will refer to these analyses, neglecting subject-wise information, as first-level analyses (and to those based on subject-wise performance measures as second-level analyses). Thereby, we avoid the per-subject lesion load bias that is introduced when one score is calculated per image. For example, missing a small lesion in an image with only this missed lesion (DSC=0) would have more weight than missing a similar lesion in an image with many other detected lesions (DSC>0).

We further investigate whether the performance of lesion segmentation varies across brain regions to identify the drivers of the metric values and possible location-dependent variabilities of LST-AI segmentation performance. To this end, we use the location-annotated lesion maps and generate binary lesion maps for each region by only selecting lesion voxels labeled as part of the corresponding region. Using the above evaluation metrics, first-level analysis is conducted for each region and results from different regions and the whole brain are compared to each other.

### 2.4.2. Lesion detection

In addition to the previous metrics, which quantify the accuracy of lesion segmentation at the voxel level, it is important to evaluate lesion segmentation methods with regard to their ability to detect lesions. In particular, this aspect is crucial in MS, since its diagnosis relies on the detection of lesions (and not on the exact measurement of their volume). To this end, we extract the following scores from the animaSegPerfAnalyzer tool: SensL, the lesion detection sensitivity; PPVL, the positive predictive value for lesions; F1 score, a metric which considers both lesion detection sensitivity and positive predictive value for lesions. SensL and PPVL are calculated according to equations (3) and (2), respectively (on the lesion level rather than on the voxel level). The F1 score is calculated as follows:

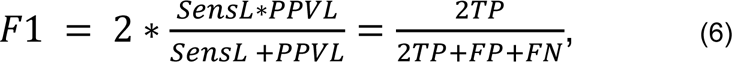

which is equal to the equation (1) and can therefore be considered as a lesion-wise DSC.

The anima evaluation toolbox also offers the animaDetectedComponents tool that can be used to investigate the detection of each lesion individually. For each image, the tool generates a list with lesions that are present in the manually segmented lesion map. It indicates, for each lesion, the volume in the manually segmented lesion map and whether it was detected by the automated segmentation method. This enables the assessment of the increase or decrease of lesion detection in relation to lesion volumes. Both tools (animaSegPerfAnalyzer and animaDetectedComponents) consider a lesion in the manual segmentation as detected if it overlaps with at least 10% with the lesion voxels in the automatically generated lesion map.

## 3. Results

We evaluate LST-AI in multiple aspects; we report both voxel-wise and lesion-wise scores, as both volume and number are established measures of lesion load. We start with lesion segmentation (3.1) across the whole brain and across subjects (second-level analyses). We then report the performance across lesions (first-level analyses) both across brain regions (3.2) and in relation to lesion size (3.3).

### 3.1. Second-level lesion segmentation across the whole brain

Lesion segmentation evaluation is conducted across all datasets as well as for each dataset individually. An overview of the results of each segmentation method across all datasets is provided in Table 3. A table with all anima metrics and results per case is included in the supplementary material. In Figure 5, we present the lesion maps (of subject 05 of the msljub dataset) of the different segmentation methods applied in this study.

**Figure 5.**
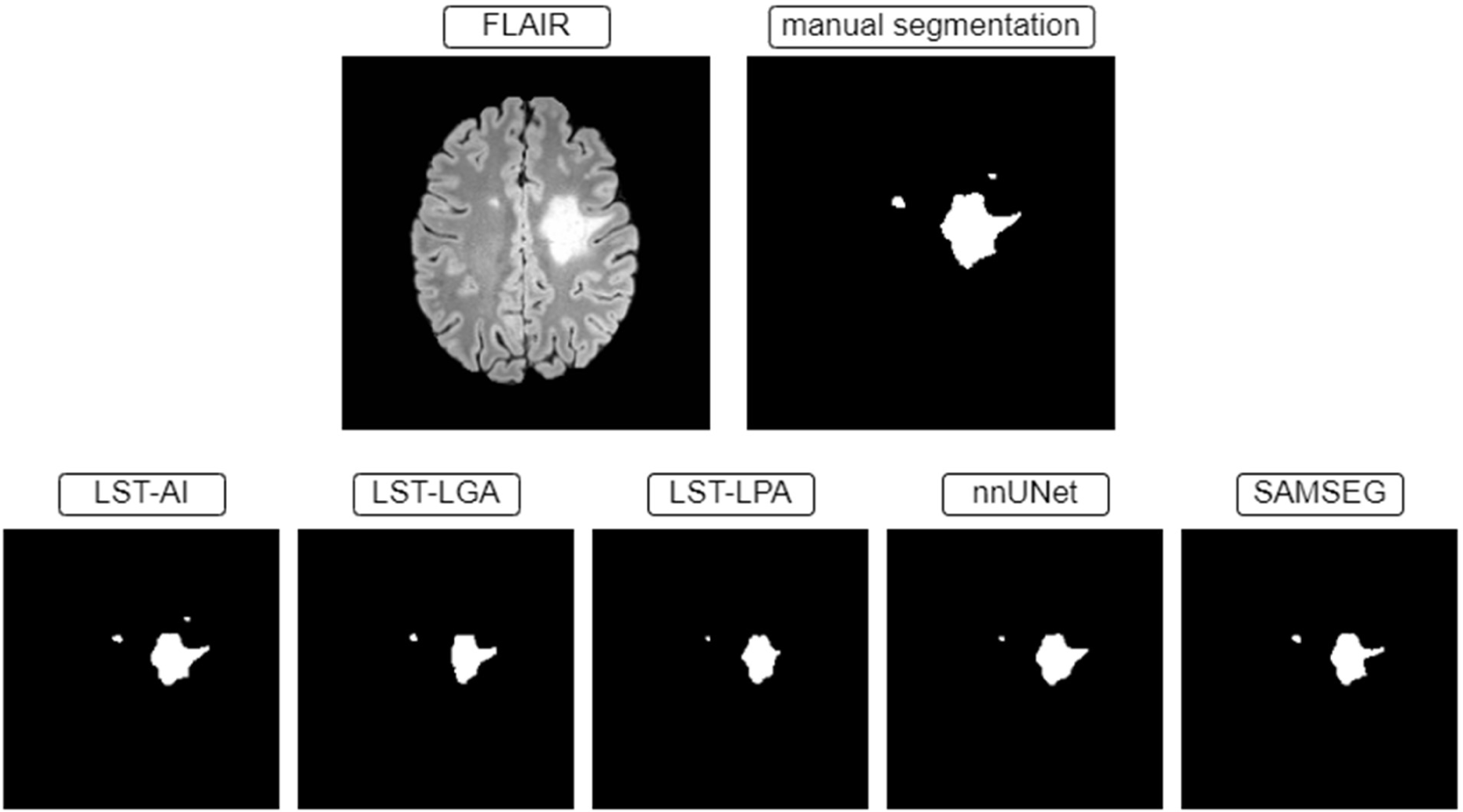
Binary lesion maps generated by the different lesion segmentation methods applied in this study. As reference, the first row shows the underlying FLAIR image as well as the manual segmentation (which is the ground truth). Each method provides slightly different lesion maps, and, in the slice presented here, only LST-AI detects all lesions present in the ground truth.

**Table 3.** The results of the lesion segmentation evaluation (second-level analysis across all test datasets) of each segmentation tool are presented. The metrics were calculated for each image in the test datasets, and values were subsequently averaged across all images. The averages are reported as mean +/- standard deviation. Abbreviations: ASD: average surface distance, DSC: dice similarity coefficient, PPV: positive predictive value, PPVL: lesion-wise positive predictive value, SensL: lesion-wise sensitivity

|  | voxel-wise |  |  |  | lesion-wise |  |  |
| --- | --- | --- | --- | --- | --- | --- | --- |
| tool | DSC | PPV | sensitivity | ASD | F1 | SensL | PPVL |
| **LST-AI** | 0.67 +/- 0.14 | 0.73 +/- 0.15 | 0.66 +/- 0.17 | 0.37 +/- 1.12 | 0.63 +/- 0.15 | 0.70 +/- 0.19 | 0.64 +/- 0.20 |
| **LST-LGA** | 0.42 +/- 0.22 | 0.80 +/- 0.21 | 0.32 +/- 0.20 | 1.43 +/- 2.81 | 0.22 +/- 0.15 | 0.20 +/- 0.14 | 0.41 +/- 0.26 |
| **LST-LPA** | 0.44 +/- 0.22 | 0.79 +/- 0.15 | 0.34 +/- 0.20 | 1.35 +/- 2.21 | 0.23 +/- 0.14 | 0.25 +/- 0.15 | 0.34 +/- 0.23 |
| **nnUNet** | 0.51 +/- 0.20 | 0.90 +/- 0.07 | 0.38 +/- 0.18 | 1.36 +/- 4.18 | 0.46 +/- 0.19 | 0.40 +/- 0.21 | 0.64 +/- 0.21 |
| **SAMSEG** | 0.55 +/- 0.20 | 0.72 +/- 0.21 | 0.49 +/- 0.19 | 1.46 +/- 4.57 | 0.38 +/- 0.18 | 0.32 +/- 0.18 | 0.57 +/- 0.21 |

The proposed method outperforms the benchmark methods in all categories except for PPV and PPVL. LST-LGA, LST-LPA, and the nnUNet yield higher PPV values (PPV=0.79-0.90) than LST-AI (PPV=0.73), and only the nnUNet yields a PPVL value as high as LST-AI (PPVL=0.64). Notably, LST-AI achieves higher DSC and F1 scores (DSC=0.67 +/- 0.14; F1=0.63 +/- 0.15) compared to the other methods (DSC=0.42-0.55; F1=0.22-0.46), indicating superior segmentation performance both on a voxel-wise and on a lesion-wise level. The lowest ASD is also obtained with LST-AI, indicating more accurate lesion contouring compared to the benchmark methods. Overall the results show that LST-AI is able to identify more true lesions while increasing the fraction of correctly identified lesions among all segmented lesions compared to the benchmark methods.

Evaluating datasets individually (Table 4), we observe the most variability across datasets in ASD.

**Table 4.** The results of the LST-AI lesion segmentation evaluation (second-level analysis) of each test dataset are presented. The metrics were calculated for each image in the respective test dataset, and values were subsequently averaged across all images. The averages are reported as mean +/- standard deviation. Abbreviations: ASD: average surface distance, DSC: dice similarity coefficient, PPV: positive predictive value, PPVL: lesion-wise positive predictive value, SensL: lesion-wise sensitivity

|  | voxel-wise |  |  |  | lesion-wise |  |  |
| --- | --- | --- | --- | --- | --- | --- | --- |
| dataset | DSC | PPV | sensitivity | ASD | F1 | SensL | PPVL |
| All datasets<br>n=103 | 0.67 +/- 0.14 | 0.73 +/- 0.15 | 0.66 +/- 0.17 | 0.37 +/- 1.12 | 0.63 +/- 0.15 | 0.70 +/- 0.19 | 0.64 +/- 0.20 |
| msisbi<br>n=21 | 0.61 +/- 0.13 | 0.72 +/- 0.11 | 0.54 +/- 0.15 | 0.41 +/- 0.66 | 0.57 +/- 0.12 | 0.55 +/- 0.15 | 0.61 +/- 0.13 |
| msljob<br>n=30 | 0.74 +/- 0.10 | 0.80 +/- 0.07 | 0.70 +/- 0.14 | 0.21 +/- 0.88 | 0.70 +/- 0.10 | 0.62 +/- 0.13 | 0.83 +/- 0.11 |
| mssgtest<br>n=37 | 0.65 +/- 0.16 | 0.68 +/- 0.19 | 0.68 +/- 0.16 | 0.59 +/- 1.60 | 0.63 +/- 0.17 | 0.83 +/- 0.14 | 0.55 +/- 0.22 |
| mssegtrain<br>n=15 | 0.67 +/- 0.16 | 0.72 +/- 0.16 | 0.67 +/- 0.19 | 0.12 +/- 0.24 | 0.61 +/- 0.15 | 0.77 +/- 0.23 | 0.53 +/- 0.09 |

### 3.2. First-level segmentation across brain regions

In the PV region, LST-AI shows slightly higher first-level DSC scores than the other methods. The difference in terms of first level DSC scores is more pronounced in the other three regions, with only LST-AI reaching DSC >0.47 (other methods: DSC=0.03-0.31). Similarly, the highest first-level DSC score within the whole brain is obtained with LST-AI. The results of the different lesion segmentation methods are presented in Table 5 and Figure 6.

**Figure 6.**
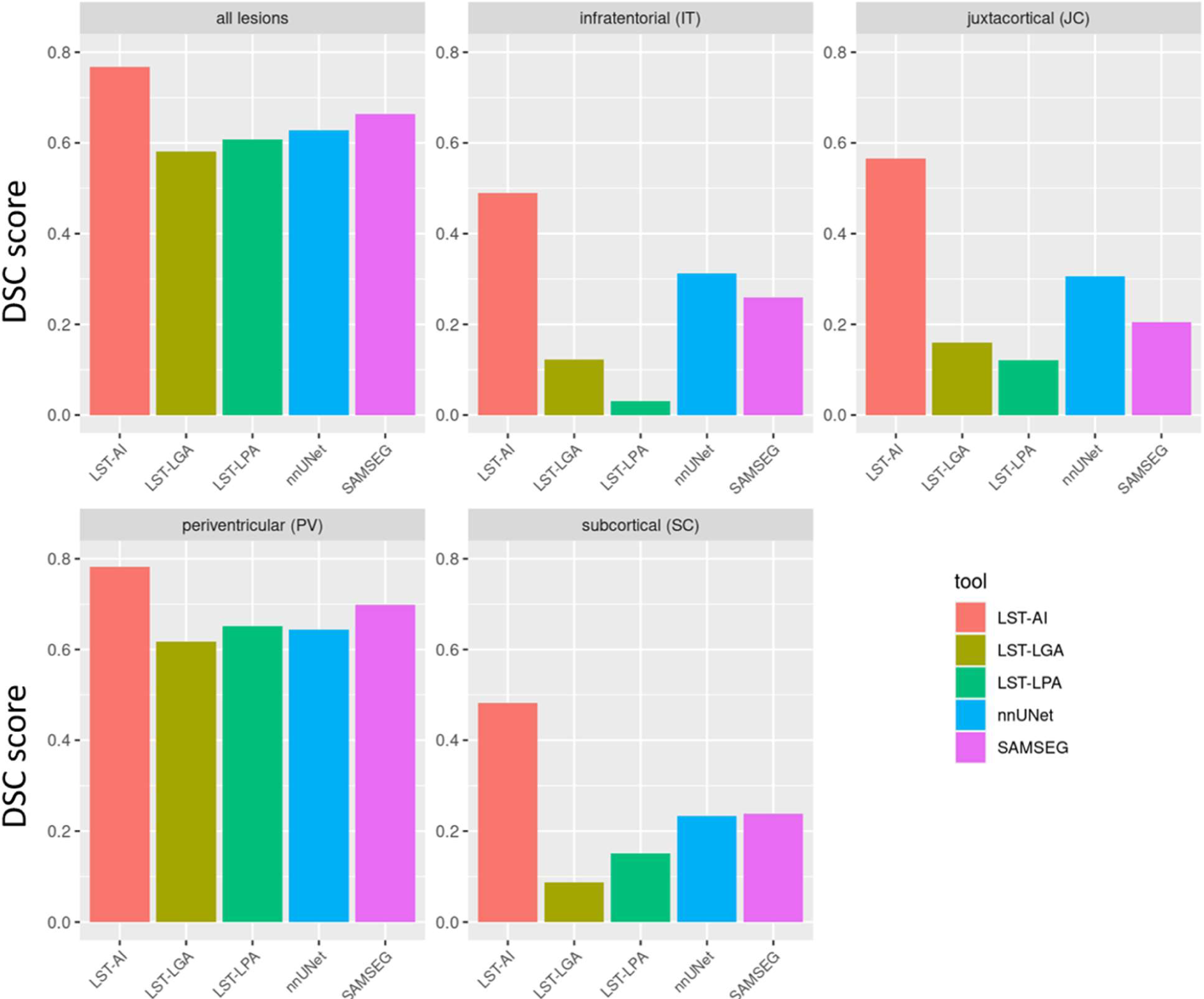
First-level DSC scores (across all test datasets) of each lesion segmentation tool are provided for lesions in different brain regions: all lesions in the whole brain, infratentorial lesions, juxtacortical lesions, periventricular lesions, and subcortical lesions.

**Table 5.** The first-level DSC score (across all test datasets) of each segmentation tool in different brain regions are presented in this table.

| tool | Periventricular (PV) | Infratentorial (IT) | Juxtacortical (JC) | Subcortical (SC) | Whole brain |
| --- | --- | --- | --- | --- | --- |
| **LST-AI** | 0.78 | 0.49 | 0.57 | 0.48 | 0.77 |
| **LST-LGA** | 0.62 | 0.12 | 0.16 | 0.09 | 0.58 |
| **LST-LPA** | 0.65 | 0.03 | 0.12 | 0.15 | 0.61 |
| **nnUNet** | 0.64 | 0.31 | 0.31 | 0.23 | 0.63 |
| **SAMSEG** | 0.70 | 0.26 | 0.21 | 0.24 | 0.66 |

### 3.3. First-level lesion detection in relation to lesion size

The lesion volume distribution of the test set is illustrated in Figure 7 (bin width of 10mm^3^). The distribution shows a fast and steep decline with the most frequent lesions being small. This is critical as there is no commonly accepted minimum lesion volume (Grahl et al., 2019); moreover, accurate manual lesion segmentation is challenging, cumbersome, and sometimes overwhelming, even for expert readers. In Figure 8, we illustrate the accuracy of lesion detection in relation to lesion volume. For this, we divided lesions into groups according to their size: 1-10 mm^3^, 11-100 mm^3^, 101-1000 mm^3^, 1001-10000 mm^3^, and larger than 10000 mm^3^. Small lesions (< 10 mm^3^) are detected worse. With increasing lesion size, the detection rate increases for all methods, with LST-AI showing the steepest incline. Hence, the advantage of LST-AI also applies to small lesions. Notably, the overall performance scores are considerably better for lesions > 10mm^3^ than suggested by mere SensL scores.

**Figure 7.**
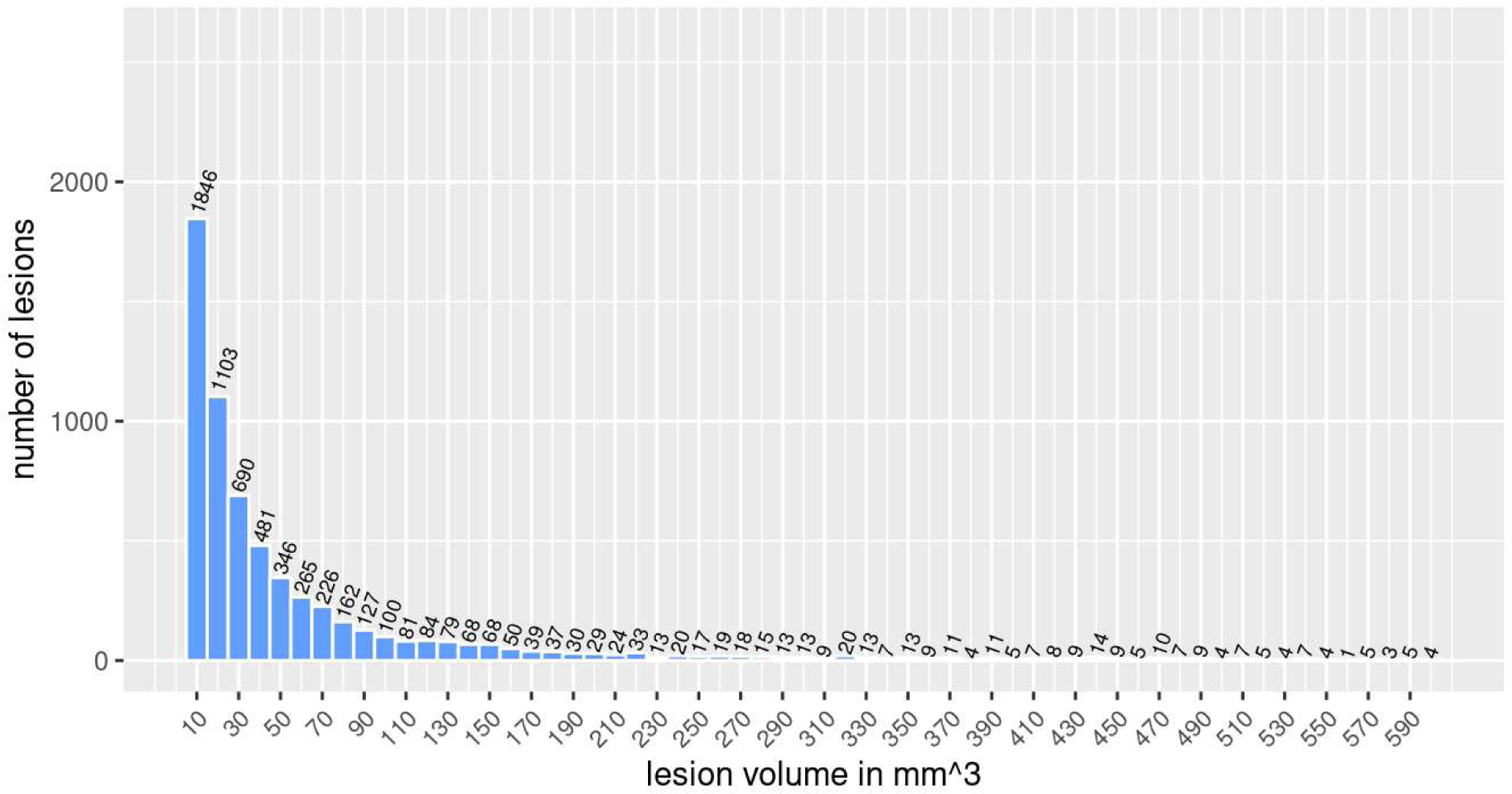
This graph shows the distribution of lesions per volume. The bars and numbers indicate how many lesions are in each volume group. We divided the lesions into groups with a volume range of 10mm^3^ and the first bar from the left shows the number of lesions with a volume between 1mm^3^ and 10mm^3^.

**Figure 8.**
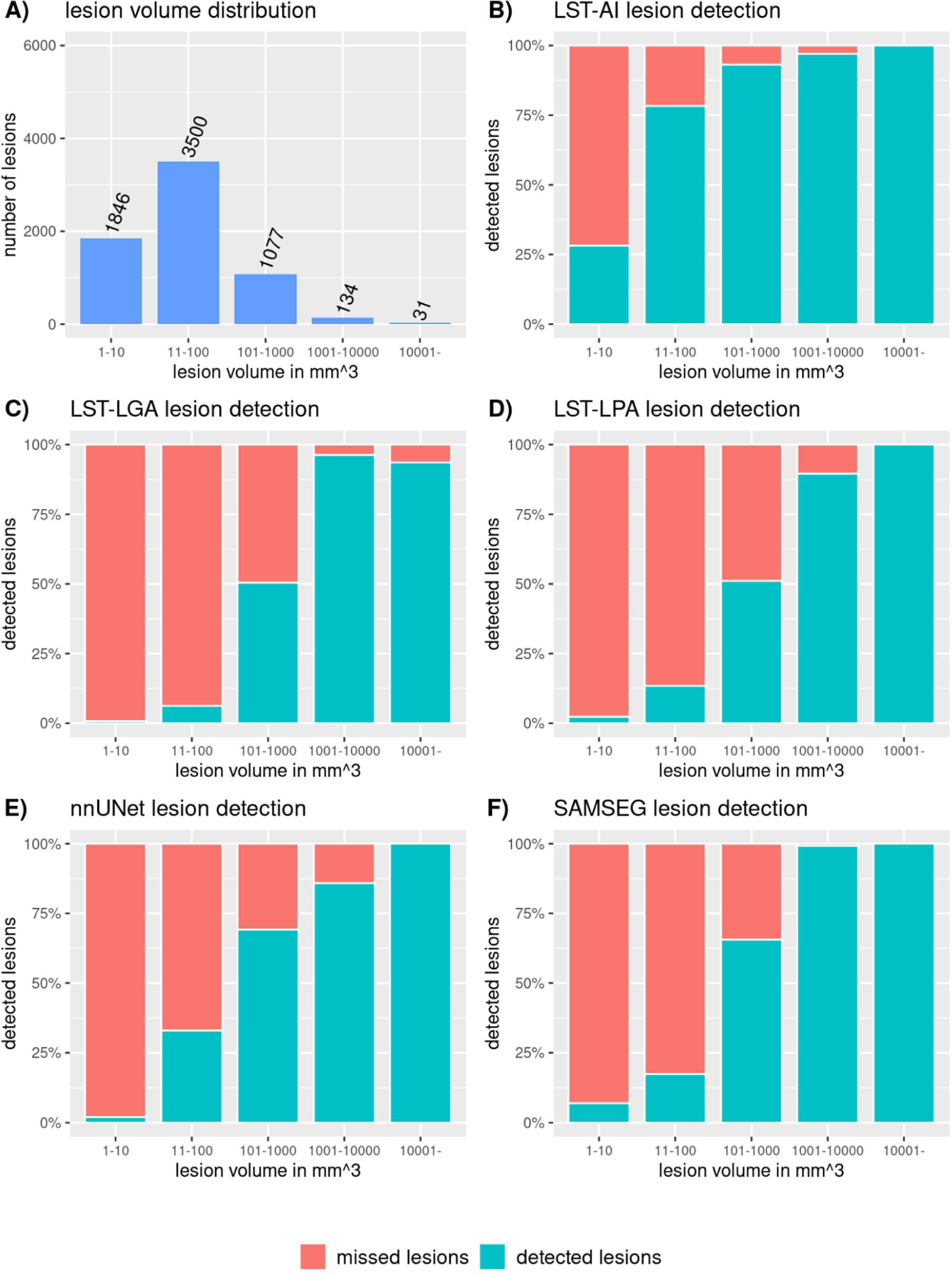
These graphs illustrate the proportion of lesions that are detected in each volume group. We divided the lesions into groups according to their volume (on the logarithmic scale): 1-10 mm^3^, 11-100 mm^3^, 101-1000 mm^3^, 1001-10000 mm^3^, and larger than 10000 mm^3^. A) shows the number of lesions distribution across the volume groups; B) - F) show the lesion detection ratios of LST-AI, LST-LGA, LST-LPA, nnUNet, and SAMSEG for the different lesion volumes. Note, how the detection rate increases with increasing lesion volume for each segmentation, whereby LST-AI yields the highest detection rates. The detection rate is given in %.

## 4. Discussion

We propose LST-AI, a new deep learning-based segmentation method for white-matter lesions in MS. It is built from an ensemble of three 3D UNets. Using LST-AI and a pair of T1w and FLAIR MRI images as input, it is possible to accurately segment lesions. We analyze the segmentation performance on multiple external datasets, thereby showing that LST-AI generalizes to data from different centers and scanners without retraining. We also compare our method to benchmark methods for validation and find excellent lesion segmentation performance of our method. In addition, LST-AI can label lesions according to their location, thereby providing further possibilities for lesion characterization in MS.

LST-AI is pre-trained on an in-house dataset consisting of 491 images and does not need to be retrained before it is applied to new data. This makes it possible to use the tool even in smaller centers, where data is scarce and only small cohorts are available. Valverde et al., 2019, have previously optimized retraining on small datasets, as their tool only requires a single case to adapt their model to new datasets. They also validated their method on the ISBI 2015 test dataset and achieved a mean DSC of 0.58 (Valverde et al., 2019). In general, high-performing segmentation models in the ISBI 2015 challenge were CNN-based (trained on ISBI 2015 training dataset) and reported DSC scores ranging between 0.50 and 0.68 (Ma et al., 2022; Zhang & Oguz, 2021). However, assessing generalizability of segmentation models requires validation on external datasets. This has been done in recent studies, which used different train and test set pairings, including in-house and publicly available data such as ISBI 2015 and MICCAI 2016 data (e.g., train on in-house data and test on MICCAI 2016 data) (Billot et al., 2021; Cerri et al., 2021; Gentile et al., 2023; Kamraoui et al., 2022; X. Li et al., 2022; McKinley et al., 2021; Rakić et al., 2021). Overall, using train and test sets from different image domains led to lower and more variable DSC scores. For example, in the study by Kamraoui et al. (2022), the segmentation performance on the ISBI 2015 test dataset drops when models are trained on in-house data (DSC=0.13-0.48) compared to when they are trained on the ISBI training dataset (DSC=0.64-0.67). On the MICCAI 2016 dataset, however, the models trained on the in-house training dataset showed robust and high DSC scores (0.65-0.72) (Kamraoui et al., 2022). This highlights the impact of differing image domains in train and test sets and the need for validation on multiple test datasets, which can provide a more realistic representation of a model’s generalizability. In this study, image domain heterogeneity is simulated by the validation of our method on multiple datasets, which were also part of MS lesion segmentation challenges of the ISBI 2015 conference and the MICCAI 2016 conference (Carass et al., 2017; Commowick et al., 2018, 2021). While our model achieves similar scores (mean DSC of 0.61 and 0.65 for ISBI 2015 and MICCAI 2016, respectively) as the top-performing models in both challenges, we want to emphasize that, in contrast to the participating models, our model is not specifically trained on the corresponding training datasets provided in the challenges. These two scores are also close to the inter-rater DSC scores of the expert segmentation used in the challenges (DSC of 0.63 and 0.66-0.76 in ISBI 2015 and MICCAI 2016, respectively) (Carass et al., 2017; Commowick et al., 2021). Other studies investigating the generalizability of their model on external data reported similar DSC scores in the range of 0.48 - 0.72 (Cerri et al., 2021; Kamraoui et al., 2022; McKinley et al., 2021; Rakić et al., 2021). Regarding LST-AI, the DSC scores for the three external datasets (range: 0.61-0.74) underline the good generalization of our model and its reliable application to multicenter data acquired with different scanners and protocols. Overall, results from both second- and first-level analysis show high segmentation performance of LST-AI on unseen data. In contrast, the lower performance of the other methods, e.g., the pre-trained nnUNet, suggests the need for adaptation of these methods through retraining. We believe that using an ensemble approach including multiple pre-trained UNets translates into robustness against performance variability of individual 3D UNets and, therefore, generalizes better across different imaging protocols and centers. Of note, the mean PPV and PPVL values of the benchmark methods are comparable to those of LST-AI. However, this appears to happen at the expense of sensitivity, where LST-AI clearly outperforms the other methods at the voxel and lesion level. Compared to the literature, lesion-wise sensitivity of LST-AI on MICCAI 2016 data (SensL=0.83) and ISBI 2015 data (SensL=0.55) is in the same range as previously reported values (Carass et al., 2017; Commowick et al., 2018; Kamraoui et al., 2022; Krishnan et al., 2023; Ma et al., 2022; Zhang & Oguz, 2021). With regard to clinical applicability of automated lesion segmentation tools, the sensitivity is crucial as diagnosing and monitoring MS relies on the detection of (new) lesions. A newly published method, namely BIANCA-MS (Gentile et al., 2023), has also been validated using the MICCAI 2016 test dataset and yielded results similar to ours in terms of DSC and false positives (in terms of lesion detection). However, the median number of false negatives was equal to 11(IQR: 18) for BIANCA-MS, whereas LST-AI yields a median number of false negatives equal to 4 (IQR: 8), again highlighting the high sensitivity of our proposed method towards lesion detection.

In MS, lesion location within the brain may play an important role in identifying different disease patterns (Pongratz et al., 2023). In the LST-AI toolbox, a method is included which is able to classify lesions into four categories according to their location (PV, IT, JC, and SC). This makes it possible to seamlessly analyze the lesion load in different brain regions relevant to MS. When looking at the segmentation performance in the four different brain regions, it stands out that, among all methods included in this publication, LST-AI shows the highest (first-level) DSC score in all regions. The increased lesion segmentation performance in the JC region is a particularly relevant finding, since segmentation of lesions close to the cortex based on T1w and FLAIR images has always been a challenge in MS. Also, juxtacortical lesions are thought to be very specific for MS and are strongly associated with clinical disability (Calabrese et al., 2012), making their detection very important.

We also investigated the lesion detection in relation to lesion volume and we found that LST-AI has a higher lesion detection sensitivity for small lesions than the benchmark methods. Similar to previous reports by Commowick et al. (2018) and Rakić et al. (2021), we also found that it is particularly hard to detect small lesions (<10mm^3^). Nonetheless, the steep incline of lesion detection with lesion size provides a promising perspective for the integration of automated lesion segmentation tools in clinical settings, since it can help clinicians to detect lesions faster and to diagnose and monitor MS more accurately.

Our study does not come without limitations. First, our model requires T1w and FLAIR image pairs, which might not always be available. Second, although less pronounced than in the benchmark methods, our model still shows a decrease in lesion detection efficiency with decreasing lesion volumes. Even though the explainability of features learned via CNNs and more specifically U-Nets have been comparatively well studied, they still lack some interpretability in contrast to methods leveraging manually selected features. In addition, preprocessing is included in the LST-AI toolbox and includes registration to MNI space, which ensures identical image dimensions and orientation before segmenting lesions. However, preprocessing steps are known to be crucial in segmentation tasks. Hence, exploring and applying different preprocessing steps could possibly change the performance on some datasets.

In conclusion, we introduce LST-AI, a new lesion segmentation toolbox and make it publicly available on GitHub (https://github.com/CompImg/LST-AI). It includes a preprocessing pipeline as well as an ensemble of three 3D UNets with binary cross-entropy and Tversky loss, making it a holistic lesion segmentation tool, enabling easy-to-implement, quick, and accurate automated lesion segmentation for MS research without retraining and fine-tuning. We validated its robustness on multiple datasets and found excellent performance. We believe that, in future studies, LST-AI should replace LST.

## Data Availability

All data produced in the present study are available upon reasonable request to the authors

## Acknowledgments

We thank Naga Karthik Enamundram and Joshua Newton for helpful discussions around the packaging of LST-AI, the evaluation of the different algorithms using the anima toolbox, and for visualization of the U-Net architecture.

## Data and code availability

We provide our toolbox as source code, command line tool and dockerized application at https://github.com/CompImg/LST-AI.

## Author contributions

Tun Wiltgen: Conceptualization, Methodology, Software, Formal analysis, Investigation, Data curation, Writing - original draft, Visualization

Julian McGinnis: Conceptualization, Methodology, Software, Formal analysis, Data curation, Writing - original draft, Visualization

Sarah Schlaeger: Investigation, Resources, Data curation, Writing - review & editing

Florian Kofler: Resources, Software

Cuici Voon: Investigation, Data curation, Writing - review & editing

Achim Berthele: Resources, Writing - review & editing

Daria Bischl: Resources, Data curation, Writing - review & editing

Lioba Grundl: Resources, Data curation, Writing - review & editing

Nikolaus Will: Resources, Data curation, Writing - review & editing

Marie Metz: Resources, Data curation, Writing - review & editing

David Schinz: Resources, Data curation, Writing - review & editing

Dominik Sepp: Resources, Data curation, Writing - review & editing

Philipp Prucker: Resources, Data curation, Writing - review & editing

Benita Schmitz-Koep: Resources, Data curation, Writing - review & editing

Claus Zimmer: Resources, Writing - review & editing

Bjoern Menze: Resources, Writing - review & editing

Daniel Rückert: Resources, Writing - review & editing

Bernhard Hemmer: Resources, Writing - review & editing

Jan Kirschke: Investigation, Resources, Data curation, Writing - review & editing

Mark Mühlau: Conceptualization, Methodology, Resources, Writing - review & editing

, Supervision, Project administration, Funding acquisition

Benedikt Wiestler: Conceptualization, Methodology, Software, Formal analysis, Data curation,

Writing - original draft, Supervision, Project administration, Funding acquisition

